# The relationship between antihypertensive medications and mood disorders: analysis of linked healthcare data for 1.8 million patients

**DOI:** 10.1101/19001719

**Authors:** Richard J Shaw, Daniel Mackay, Jill P. Pell, Sandosh Padmanabhan, David S. Bailey, Daniel J. Smith

## Abstract

**Background:** Recent work suggests that antihypertensive medications may be useful as repurposed treatments for mood disorders. Using large-scale linked healthcare data we investigated whether certain classes of antihypertensive, such as angiotensin antagonists and calcium channel blockers, were associated with reduced risk of new-onset Major Depressive Disorder (MDD) or bipolar disorder.

**Method:** Two cohorts of patients treated with antihypertensives were identified from Scottish prescribing (2009-2016) and hospital admission (1981-2016) records. Eligibility for cohort membership was determined by receipt of a minimum of four prescriptions for antihypertensives within a 12-month window. One treatment cohort (n=538,730) included patients with no previous history of mood disorder, whereas the other (n=262,278) included those who did. Both cohorts were matched by age, sex and area deprivation to untreated comparators. Associations between antihypertensive treatment and new-onset MDD or bipolar episodes were investigated using Cox regression.

**Results:** For patients without a history of mood disorder, antihypertensives were associated with increased risk of new-onset MDD. For angiotensin antagonist monotherapy, the hazard ratio (HR) for new-onset MDD was 1.17 (95%CI 1.04-1.31). Beta blockers’ association was stronger (HR 2.68; 95%CI 2.45-2.92), possibly indicating pre-existing anxiety. Some classes of antihypertensive were associated with protection against bipolar disorder, particularly angiotensin antagonists (HR 0.46; 95%CI 0.30-0.70). For patients with a past history of mood disorders, all classes of antihypertensives were associated with increased risk of future episodes of MDD.

**Conclusions:** There was no evidence that antihypertensive medications prevented new episodes of MDD but angiotensin antagonists may represent a novel treatment avenue for bipolar disorder.

## Introduction

Major depressive disorder (MDD) and bipolar disorder (BD) are leading causes of disability globally (Ferrari et al., 2016; Vos et al., 2016). More than one third of MDD patients will not respond to first-line antidepressants (Linde et al., 2015) and BD is challenging to treat, with antidepressants ineffective for most patients (Sidor & Macqueen, 2011). In recent years there has been little progress in the development of new medications for mood disorders (except perhaps for ketamine in depression; Krystal, Abdallah, Sanacora, Charney, & Duman, 2019) but there is currently considerable interest in the possibility of repurposing medications from other areas of medicine.

Specifically, it has been suggested that certain classes of antihypertensive medication (particularly calcium-channel blockers, CCBs and angiotensin antagonists, AA) may have a role as repurposed treatments for MDD or BD (Harrison et al., 2016; Harrison, Tunbridge, Dolphin, & Hall, 2019; Saavedra, 2017; Vian et al., 2017).

To date, the evidence for repurposing antihypertensive drugs to treat MDD is limited (Chowdhury, Berk, Nelson, Wing, & Reid, 2019; Vian et al., 2017). AAs have been reported in small observational studies to be associated with better mental health outcomes (Ahola, Harjutsalo, Forsblom, & Groop, 2014; Boal et al., 2016; Brownstein et al., 2018; Johansen, Holmen, Stewart, & Bjerkeset, 2012; Nasr, Crayton, Agarwal, Wendt, & Kora, 2011; Williams et al., 2016). However, a recent large linkage study found that although initial prescriptions for AAs were associated with increased risk of depression and bipolar disorder, people receiving longer-term prescriptions for AAs were not at increased risk (Lars Vedel Kessing et al., 2019; L. V. Kessing et al., 2019).

Since the 1980s, CCBs such as verapamil have been suggested as possible treatments for mania (Celano et al., 2011). This was not supported by a review of six double-blind randomised studies and 17 observational studies which found that no evidence for efficacy of CCBs in mania (Cipriani et al., 2016). Nonetheless, despite the relatively limited evidence base to date, CCBs remain candidates for repurposing in BD because of their biological plausibility (Cipriani et al., 2016).

There is very little published work supporting the repurposing of other antihypertensive drug classes for MDD or BD. Case reports and some clinical trials have found that beta-blockers (BB) (particularly propranolol) may be associated with increased depressive features (Luijendijk & Koolman, 2012; Verbeek, van Riezen, de Boer, van Melle, & de Jonge) (although recent observational work suggests that BBs have little influence on mood disorder outcomes; Boal et al., 2016; Johansen et al., 2012; Ko et al., 2002; Luijendijk & Koolman, 2012; Nasr et al., 2011; Ranchord, Spertus, Buchanan, Gosch, & Chan, 2016; Verbeek et al). It is possible that depressogenic effects are restricted to more lipophilic BB (such as propranolol) which cross the blood-brain barrier (Thiessen, Wallace, Blackburn, Wilson, & Bergman, 1990; Verbeek et al.). For other antihypertensive drugs, such as diuretics and centrally acting agents, almost no evidence of any influence on mood disorder outcomes has so far been described (Celano et al., 2011; Coyne, Davis, French, & Hill, 2002; J. F. Hayes et al., 2019; Huffman & Stern, 2007; Nasr et al., 2011; Williams et al., 2016).

Our primary goal was to use Scottish national-level routine healthcare data on over 1.8 million individuals (representing more than 6 million person-years of follow-up) to assess whether patients treated with specific classes of antihypertensive medication as monotherapy were less likely to experience new-onset mood disorder episodes.

## Methods

### Data Sources

Within the National Services Scotland Safehaven, we created two datasets of cohorts and comparison groups by linking data from: the Community Health Index; Scottish Morbidity Records (SMR) datasets including SMR00 (Outpatient Attendance) from 1997 to 2016, SMR01 (General/Acute Inpatient and Day Case) from 1981 to 2016, and SMR04 (Mental Health Inpatient and Day Cases) from 1981 to 2016; the Prescribing Information System (PIS) from 2009 to 2016; and the National Records of Scotland death certificates from 1981 to 2016. Cohort 1 included patients treated with new-onset antihypertensive treatment (defined below) who had no previous record of mood disorder. Cohort 2 included patients with new-onset antihypertensive treatment plus a past record of mood disorder. Ethical approval for the project was obtained from the Public Benefit and Privacy Panel at National Health Services Scotland Information and Statistics division.

Time periods in which potential participants were eligible for inclusion were defined on the basis of prescriptions for antihypertensive medications, as well as prescriptions and hospital admission records for psychiatric disorders. To define the cohorts, we initially used PIS data from January 2009 to December 2016 to identify individuals who had a minimum of 4 prescriptions for antihypertensives (defined using British National Formulary (BNF)(Joint Formulary Committee, 2019) sections and paragraphs 2.2, 2.4, 2.5.5, and 2.6.2, see supplementary Table S1 for more details on the BNF classifications) within a window of up to 12 months, preceded by 6 months of no antihypertensive treatment record. We used receiving a minimum of 4 or more prescriptions from a single antihypertensive treatment as an inclusion criteria because typically a single prescription would cover a period of three months so a minimum of 4 prescriptions would be required to cover a period of one year. The 6 months without antihypertensive treatment were to ensure that subsequent antihypertensive prescriptions were for a *new* treatment, rather than part of an ongoing treatment regime. Patients were then excluded from cohort 1 and included in cohort 2 if they had been prescribed psychiatric medication (indicated by BNF sections 4.1, 4.2, 4.3) within the same window. Following that, we also excluded people from cohort 1 and included them in cohort 2 if they had been admitted to hospital for psychiatric treatment (as indicated by a clinic appointment with the general psychiatry speciality in SMR00 database, ICD10 codes F10-F48, X60-X84 and Z91.5 and ICD9 codes 290-301 and 303-305 in the SMR01 database and a record in the SMR04 databases) during the antihypertensive treatment window and for the preceding 10 years. At this stage, the number of potential cohort 1 members was 968,930 for cohort 1 (See supplementary Figure S1), and for potential cohort 2 members this number was 555,975 (See supplementary Figure S2).

Given that the pool of people for the comparison group was limited, for both cohorts we selected individuals who had an eligible period that ended after 31/12/2009. We aimed to match cohort members with comparisons using a 1:1 ratio for cohort 1 and 1:2 for cohort 2. Comparisons for each cohort were initially selected on the criteria that they had received no antihypertensive medication between 2009 and 2016, and then on the same criteria as their corresponding cohort with respect to psychiatric treatment. Matching was on the basis of age (+/- 2 years), sex and Scottish Index of Multiple Deprivation. Subsequently, cohort-comparison pairs were excluded from analysis if they either had a death record prior to the end of their eligible treatment window, or were outside the age range of 18 to 100. This resulted in 538,789 cohort-comparison pairs for cohort 1, and 272,278 cohort 2 members matched to 502,937 comparators. Descriptive statistics for unmatched but otherwise eligible patients are shown in supplementary file Table S2 for Cohort 1 and Table S3 for Cohort 2. The main barrier to matching cohort members was age, with it being much harder to find matches for older cohort members particularly for cohort 2 for which we used a higher matching ratio.

### Antihypertensive monotherapy and polytherapy status

Participants were identified for treatment on the basis of prescribing records for antihypertensive medication within the 2009-2016 PIS dataset. These codes were then used to classify participants into specific classes on the basis of the prescription of thiazide diuretics, beta-blockers (BB), angiotensin antagonists (AA) (including ACE-inhibitors, angiotensin receptor blockers (ARB) and renin inhibitors), and calcium channel blockers (CCBs). Patients were included in antihypertensive monotherapy groups if during the 12 months prior to the end date of the eligibility period they received 4 or more prescriptions from a single class of antihypertensive treatment and no prescriptions for any of the other classes. For the analysis of monotherapy, individuals were subsequently censored at the date on which they received a prescription for an antihypertensive drug outside their monotherapy class. Patients were considered to be on polytherapy if during the last three months of the eligible treatment window they received antihypertensive medication from two or more of thiazide diuretics, BBs, AA or CCBs. Remaining study participants who had received at least four antihypertensive treatments but were not eligible for the monotherapy or polytherapy groups were classified as ‘other antihypertensive.’ See Table S4 in the supplementary file for the frequency of each of the different combinations.

### Outcome measures

We had two outcome measures indicating new-onset of treatment for episodes of either MDD or BD, indicated by receipt of medications (PIS) or psychiatric admissions (SMR04) subsequent to the end date of the eligibility window used to define the cohorts. Using PIS data, new onset of treatment for MDD was identified by the prescription of any antidepressant drugs (BNF section 4.3), and new onset of BD was identified by a prescription for antipsychotics and drugs used for mania and hypomania (BNF section 4.2). Similarly, the ‘main’ and ‘other diagnoses’ fields in SMR04 were used to identify first episodes for treatment for MDD or BD (ICD10 codes F32 and F33 used to indicate MDD and ICD10 codes F30 and F31 used to indicate BD).

### Confounding variables

Medical comorbidities for study members were defined by having ever received specific prescriptions or attended hospital for specific conditions as indicated in available PIS, SMR00, SMR01 and SMR04 records, up to end date of the eligibility window.

Using PIS data, ever prescribed cardiovascular medication (other than antihypertensives) was defined using prescriptions for drugs from BNF sections or paragraphs 2.1, 2.3, 2.6.1, 2.6.3, 2.6.4, 2.9 and 2.12, and treatment for diabetes was indicated by receipt of prescriptions of antidiabetic medications (BNF chapter 6.1.2.)

Confounding variables were derived from hospital records based on specific ICD10 and ICD9 codes, as indicated in the ‘main’ and ‘other conditions’ for SMR00, SMR01 and SMR04 and ‘main’ admission and ‘other’ admission condition for SMR04. History of cardiovascular disease was defined using ICD10 codes I20-I25 (Ischaemic heart diseases), and I60-I69 (Cerebrovascular diseases), and ICD 9 codes 410-414 (Acute myocardial infarction) and 430-438 (Cerebrovascular diseases). Head injuries were identified using ICD 10 codes S02.0, S02.1, S02.7 S06, S07 and ICD9 codes 800-804 and 850-854. Substance abuse was identified using ICD codes F10 to F19, and ICD9 codes 291, 292, 303 to 305. Self-harm was identified using ICD codes X60-X84 and Z91.5 and ICD 9 codes E950 – E958.

Additional confounding variables were defined for cohort 2 who had a history mental illness or mood disorders. PIS data was used to identify people who had been prescribed the following types of pharmaceutical treatment: Hypnotics or Anxiolytics (BNF Section 4.1), antidepressants (BNF Section 4.3), or drugs used in psychoses and related disorders (BNF Section 4.2). Hospital admissions records were used to identify people who had been admitted to hospital for the following conditions: Schizophrenia and delusional disorders (ICD10 Codes F10-F19 and ICD9 codes 291-292 and 303-305), MDD (ICD10 codes F32-F33, and ICD9 codes 296.2-296.3), BD (ICD10 codes F30-F32, and ICD9 codes 296.0, 296.1, and 296.9) and personality disorders (ICD10 codes F60-F69, and ICD9 code 301).

### Data analysis

All analyses were carried out within the National Safehaven using Stata 14.0 MP. The curves for first onset of mood disorder by therapy classes are presented using cumulative distributive functions, which are calculated as 100 percent minus the Kaplan-Meier estimate (Cleves, Gould, & Marchenko, 2016). Data were analysed using Cox proportional hazards models, which were stratified by cohort and control pairs, to investigate the relationship between specific therapy classes and new-onset of MDD or BD following the end of the 12 month eligibility window, after adjustment for age and the other medication and hospital admission variables. Preliminary analyses suggested that the proportional hazards assumption was violated; consequently this was addressed using a Heaviside function (Kleinbaum & Klein, 2010) with separate hazard ratios calculated for the specific therapy classes for the following five time periods: 0 to 3 months, >3 to 6 months, >6 to 12 months, >1 year to 2 years and >2 years.

## Results

Sociodemographic characteristics for cohort 1 are shown in Table 1, and a comparison of both cohorts is shown in Table S5. The mean age of the different antihypertensive treatment groups varied, with patients on BBs having the youngest mean age of 53.4 years, while those in the ‘other antihypertensive’ group had the oldest mean age of 66.6 years. Patients receiving thiazide diuretics, BBs and other patterns of antihypertensive treatments were more likely to be women, while those receiving AAs, CCBs, and polytherapy were more likely to be men. There were small differences between the groups in terms of area deprivation, with patients receiving thiazides the most affluent and those on other medications the least affluent.

**Table 1:** Sociodemographic, medical event and history variables Cohort 1.

|  | Comparison |  | Thiazide diuretics<br>(Thiazide) |  | Beta Blockers<br>(BB) |  | Angiotensin<br>Antagonists<br>(AA) |  | Calcium<br>Channel<br>Blockers<br>(CCBs) |  | Polytherapy<br>(Poly) |  | Other<br>Antihypertensives |  |
| --- | --- | --- | --- | --- | --- | --- | --- | --- | --- | --- | --- | --- | --- | --- |
|  | n | % | n | % | n | % | n | % | n | % | n | % | n | % |
|  | 538,730 | 100 | 25,555 | 100 | 73,996 | 100 | 130,110 | 100 | 57,986 | 100 | 211,282 | 100 | 39,801 | 100 |
| **Gender** |  |  |  |  |  |  |  |  |  |  |  |  |  |  |
| Male | 285,329 | 53.0 | 7,612 | 29.8 | 32,673 | 44.2 | 78,459 | 60.3 | 31,135 | 53.7 | 119,171 | 56.4 | 16,279 | 40.9 |
| Female | 253,401 | 47.0 | 17,943 | 70.2 | 41,323 | 55.8 | 51,651 | 39.7 | 26,851 | 46.3 | 92,111 | 43.6 | 23,522 | 59.1 |
| **Survival events** |  |  |  |  |  |  |  |  |  |  |  |  |  |  |
| Treatment for depression | 94,073 | 17.5 | 6,169 | 24.1 | 19,181 | 25.9 | 25,461 | 19.6 | 9,752 | 16.8 | 46,720 | 22.1 | 9,321 | 23.4 |
| Treatment for Bipolar disorder | 11,822 | 2.2 | 688 | 2.7 | 1,251 | 1.7 | 1,816 | 1.4 | 1,143 | 2.0 | 4,341 | 2.1 | 1,753 | 4.4 |
| Died | 41,319 | 7.7 | 2,211 | 8.7 | 4,177 | 5.6 | 6,615 | 5.1 | 4,092 | 7.1 | 17,767 | 8.9 | 8,785 | 22.1 |
| Changed Therapy | na |  | 11,580 | 45.3 | 14,701 | 19.9 | 43,787 | 33.6 | 18,561 | 32.0 | na |  | na |  |
| **History of hospital treatment** |  |  |  |  |  |  |  |  |  |  |  |  |  |  |
| Cardiovascular disease | 11,523 | 2.1 | 1,149 | 4.5 | 10,293 | 13.9 | 9,452 | 7.3 | 4,358 | 7.5 | 44,495 | 21.1 | 6,209 | 15.6 |
| Substance Abuse | 6,995 | 1.3 | 237 | 0.9 | 957 | 1.3 | 1,650 | 1.3 | 806 | 1.4 | 2,822 | 1.3 | 583 | 1.5 |
| Head Injury | 15,570 | 2.9 | 410 | 1.6 | 2,185 | 3.0 | 4,054 | 3.1 | 1,374 | 2.4 | 4,794 | 2.3 | 1,003 | 2.5 |
| Self-Harm | 1,373 | 0.3 | 48 | 0.2 | 317 | 0.4 | 421 | 0.3 | 155 | 0.3 | 471 | 0.2 | 130 | 0.3 |
| **History of Prescriptions** |  |  |  |  |  |  |  |  |  |  |  |  |  |  |
| Other Cardiovascular drugs | 61,154 | 11.4 | 10,009 | 39.2 | 29,355 | 39.7 | 59,563 | 45.8 | 25,688 | 44.3 | 127,965 | 60.6 | 19,235 | 48.3 |
| Diabetic drugs | 8,372 | 1.6 | 854 | 3.3 | 2,365 | 3.2 | 20,027 | 15.4 | 2,242 | 3.9 | 23,359 | 11.1 | 2,902 | 7.3 |
| **Age** |  |  |  |  |  |  |  |  |  |  |  |  |  |  |
| Mean | 59.4 |  | 64.9 |  | 53.4 |  | 56.3 |  | 63.9 |  | 61.2 |  | 66.6 |  |
| SD | 13.2 |  | 11.4 |  | 16.6 |  | 12.0 |  | 11.9 |  | 11.2 |  | 15.5 |  |
| **SIMD 2016** |  |  |  |  |  |  |  |  |  |  |  |  |  |  |
| Mean | 5.6 |  | 5.8 |  | 5.6 |  | 5.7 |  | 5.7 |  | 5.6 |  | 5.5 |  |
| SD | 2.8 |  | 2.8 |  | 2.8 |  | 2.8 |  | 2.8 |  | 2.8 |  | 2.8 |  |

“Other Antihypertensives” were defined on the basis of treatment with a combination of thiazide diuretics, diuretics, BBs, AA and/or CCBs, but not treatment with at least two of these groups within in the last 3 months of the eligible treatment window.

Medical history and mood disorder outcomes are also shown in Table 1 for cohort 1 and in Supplementary Table S6 for cohort 2. As might be expected for the comparison group, both the percentages of people having been admitted to hospital for cardiovascular disease and receiving prescriptions for other cardiovascular medicines were lower than the equivalent figures for all the antihypertensive treatment groups. However, for the other medical history measures the comparison groups fell within the range of the antihypertensive treatment groups.

### Cumulative distribution functions

The cumulative distribution functions for new-onset of an episode of MDD by therapy class are shown in Figure 1a for cohort 1 (See Supplementary Figure S3a for cohort 2). Numbers at risk and number of failures for both the treatment cohorts are shown in Supplementary Table S7. The main difference between the two cohorts was the expected higher incidence rates of MDD episodes in the first 0-3 months for cohort 2, reflecting that (by definition) these patients already had a record of mood disorder. The comparison groups for both cohorts had the lowest risk of receiving treatment for MDD throughout the follow-up period, and patients receiving BB monotherapy had the highest risk for MDD episodes. The curve for the ‘other antihypertensive’ treatment group in cohort 1 was similar to the curve for those receiving BBs in that cohort. Among all the antihypertensive monotherapy groups, those receiving AAs had the lowest risk of new-onset MDD.

**Figure 1:**
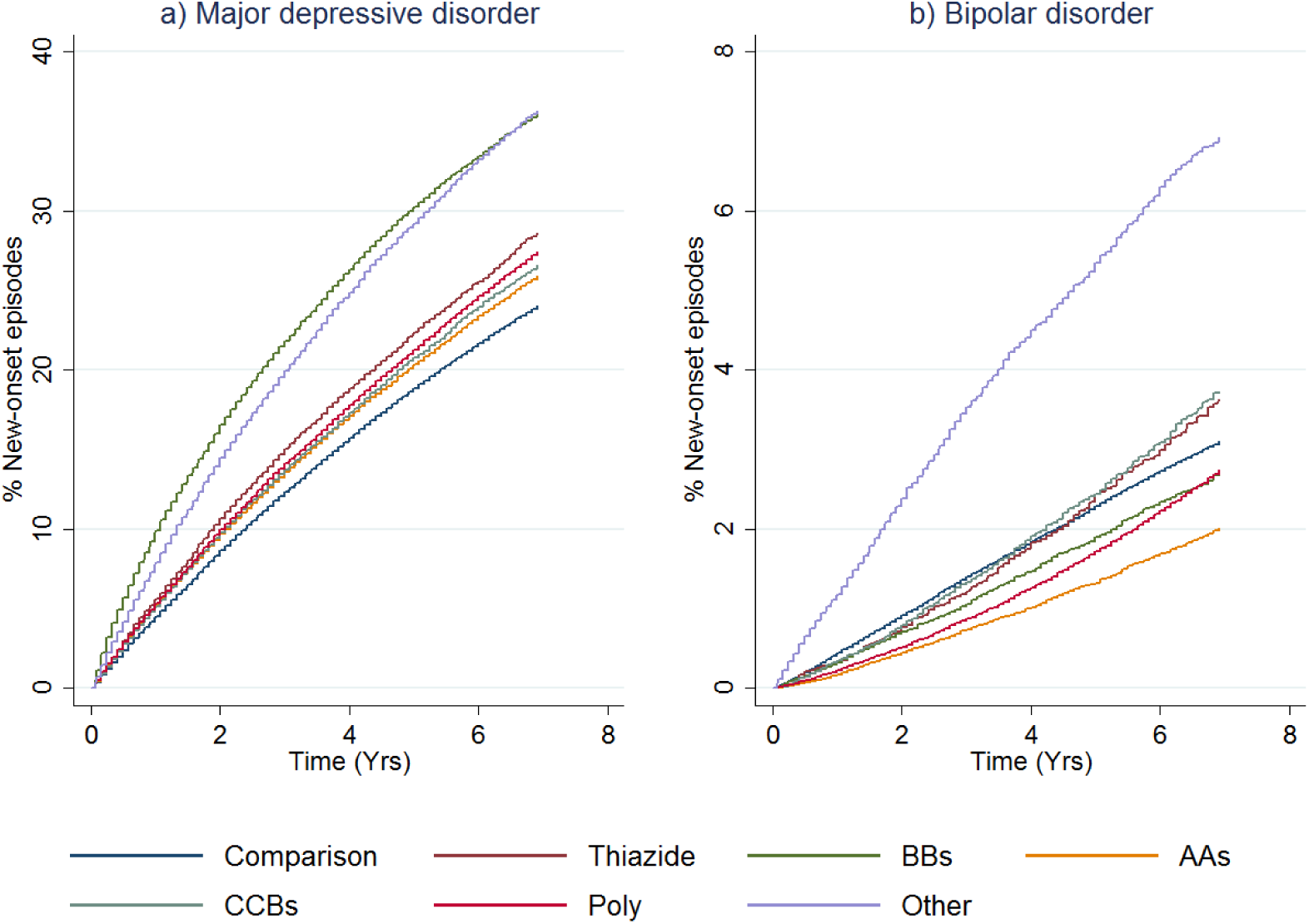
First onset of mood disorders, as indicated by receipt of prescriptions or admission to hospital, by therapy class (people without mental illness). Figure 1 shows cumulative distributions functions for major depressive disorder (panel a) and bipolar disorder (panel b) by therapy class for people without a history of treatment for mental illness (cohort 1). “Other Antihypertensives” were defined on the basis of treatment with a combination of thiazide diuretics, diuretics, BBs, AA and/or CCBs, but not treatment with at least two of these groups within in the last 3 months of the eligible treatment window.

The cumulative distribution functions for new-onset BD episodes by antihypertensive monotherapy group are shown in Figure 1b. The equivalent figure for cohort 2 is shown in Supplementary Figure S3b. For cohort 1, the ‘other antihypertensive’ group was more likely to receive treatment for new-onset BD compared to all the other classes of treatment. The comparison group curve lay between the curves for the other antihypertensive classes, and people receiving polytherapy and AAs were somewhat less likely to be treated for BD than the other therapy classes. Once the (expected) sharp incidence rate is accounted for in cohort 2, curves were similar to cohort 1.

### Cox proportional hazard models for new-onset of depression

The hazard ratios (after adjustment for hospital treatment for cardiovascular disease, substance abuse, head injury, self-harm, and pharmaceutical treatment for other cardiovascular or diabetic drugs) for the relationship between receiving a specific class of antihypertensive and new-onset MDD over time for people without a prior history of mood disorder (cohort 1) are shown in Figure 2.

**Figure 2:**
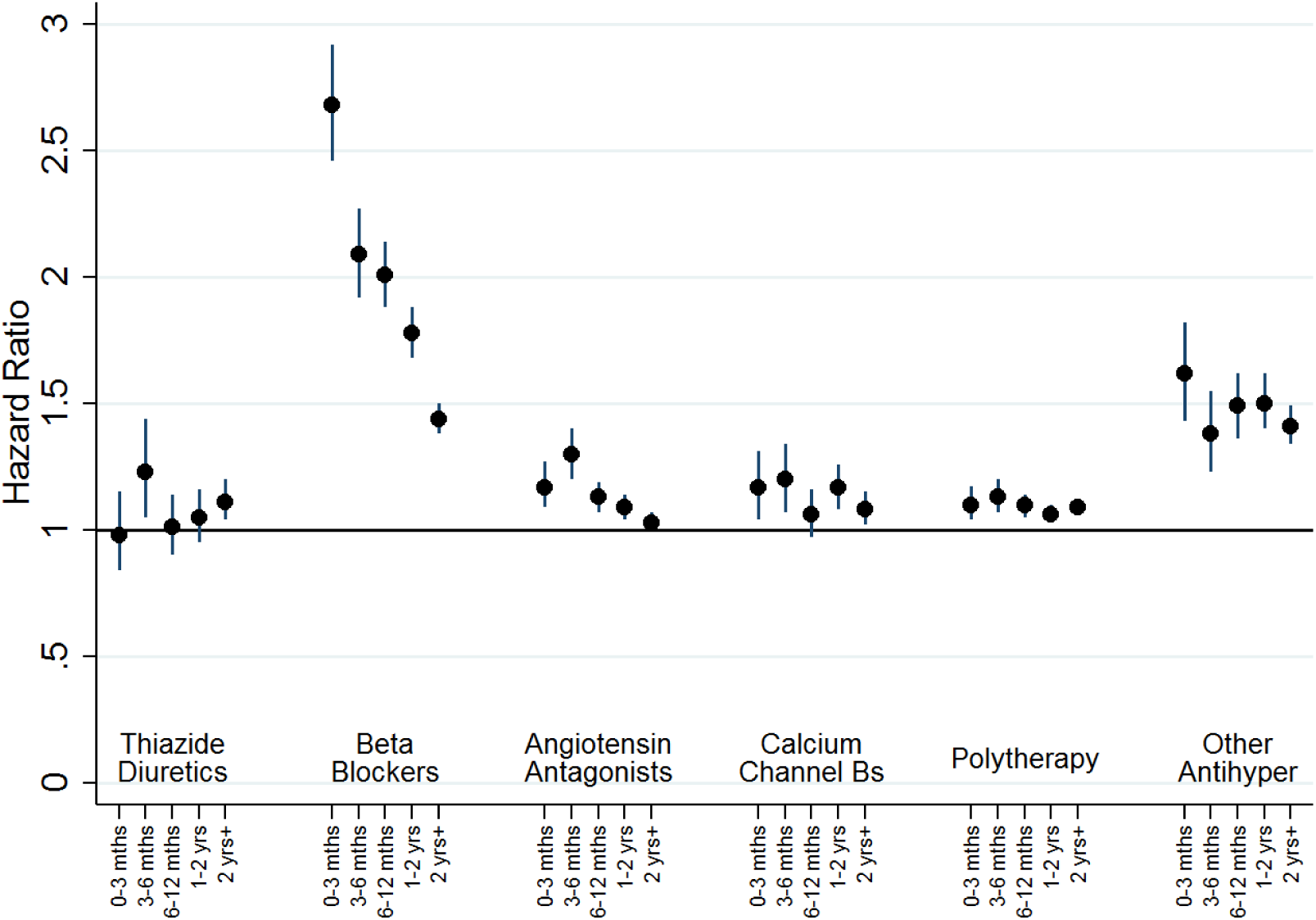
Hazard ratios for new onset depression, as indicated by receipt of prescriptions or admission to hospital, by therapy class (people without mental illness). Figure 2 shows hazard ratios for new onset depression by therapy class for people without a history of treatment for mental illness (Cohort 1). Adjustment was carried out for hospital treatment for cardiovascular disease, substance abuse, head injury, self-harm, and pharmaceutical treatment for other cardiovascular drugs and diabetic drugs. “Other Antihypertensives” were defined on the basis of treatment with a combination of thiazide diuretics, diuretics, BBs, AA and/or CCBs, but not treatment with at least two of these groups within in the last 3 months of the eligible treatment window.

For people receiving most monotherapy treatments and the polytherapy group there was a small but consistently elevated hazard ratio of around 1.2, which declined with time. For people receiving ‘other antihypertensive’ treatments, there was a slightly higher hazard ratio of around 1.5 for all time points. In contrast, the hazard ratio (HR) for people treated with BBs was initially high at 2.68 (95% Cl, 2.45-2.92) in the first three months, before declining to 2.01 (95% CI, 1.88-2.13) after 6 months, and 1.44 (95% CI, 1.38-1.50) at two years.

We explored the possibility that any associations were restricted to sub-groups among AAs and BBs During the first three months the hazard ratios for participants being treated with propranolol (N = 30,478) was 4.80 (95% CI 4.22-5.46), with this falling over time (3 to 6 months: HR 3.74, 95% CI 3.31-4.23, 6 to 12 months: HR 3.29, 95% CI 2.99-3.62, 1 to 2 years: HR 2.73, 95% CI 2.51-2.97) to just 2.04 (95% CI, 1.91-2.17) after 2 years. In contrast, the associations for atenolol (n = 16,650), other beta blockers (N = 26,868) and the two subclasses of AAs (ACE inhibitors, N = 105,064, or ARB, N = 16,071) were consistently weak, with a hazard ratio of around 1.1. We also ran additional analyses separately for men (Supplementary Figure S4) and women (Supplementary Figure S5). The odds ratios for new-onset depression for nearly all antihypertensive classes were larger for women than the corresponding odds ratios for men, however, the gender differences were small and conclusions drawn for both genders are similar.

There was greater consistency of relationships between all therapy classes of antihypertensive treatment and new-onset MDD episodes for people with a prior history of mood disorder (cohort 2; Supplementary Figure S6). For most antihypertensive groups, the hazard ratios were around 1.4 at 0-3 months and about 3 for between 3-6 months, with hazard ratios continuing to increase thereafter. However, BBs appeared to have a slightly stronger association at 0 to 3 months (HR 1.93, 95% CI 1.90-1.96) and at three to six months (HR 3.83, 95% CI 3.63-3.04). Further analyses indicated that elevated hazard ratios for people on BBs were restricted to people who had been prescribed propranolol.

### Cox proportional hazard models for new-onset bipolar disorder

In adjusted analyses for people with no previous history of mood disorder (cohort 1), most therapy classes of antihypertensive drugs were initially associated with reduced risk of BD episodes, with the risk subsequently increasing towards a null association over time (see Figure 3). The exceptions were those on BBs and other treatment groups, for whom there appeared to be some limited evidence of association with BD outcomes. As before, we investigated subgroups within AA and BBs. The associations for both ACE inhibitors and ARBs were the same as for angiotensin antagonists combined, and participants who were treated with atenolol and other BBs also had a reduced risk of being treated for BD. In contrast, those treated with propranolol had a much higher risk of being treated for BD at 0-3 months (HR 3.33, 95% CI 1.08-10.27), with the risk falling gradually thereafter. We ran separate analyses for men (Supplementary Figure S7) and women (Supplementary Figure S8) and there was little evidence of gender differences that could not have occurred by chance.

**Figure 3:**
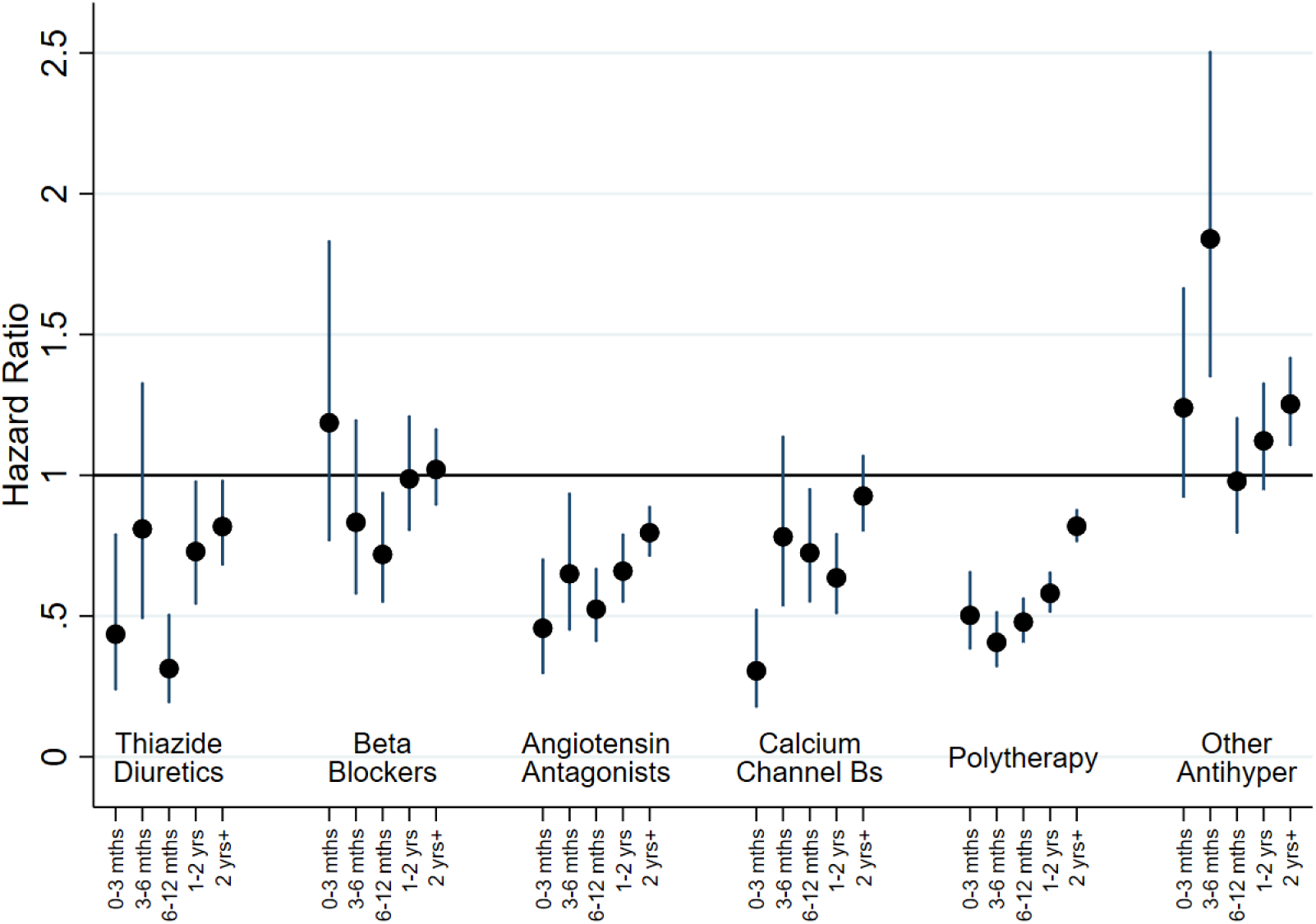
Hazard ratios for new onset bipolar disorder, as indicated by receipt of prescriptions or admission to hospital, by therapy class (people without mental illness). Figure 3 shows hazard ratios for new bipolar disorder by therapy class for people without a history of treatment for mental illness (Cohort 1). Adjustment was carried out for hospital treatment for cardiovascular disease, substance abuse, head injury, self-harm, and pharmaceutical treatment for other cardiovascular drugs and diabetic drugs. “Other Antihypertensives” were defined on the basis of treatment with a combination of thiazide diuretics, diuretics, BBs, AA and/or CCBs, but not treatment with at least two of these groups within in the last 3 months of the eligible treatment window.

A slightly more complex pattern was evident in adjusted analyses for those with a prior history of mood disorder (cohort 2; Supplementary Figure S9). Most antihypertensive classes were initially associated with a small reduction in risk during the first 3 months. However, this tended to change over time and after a year was associated with increased risk of BD. The exceptions to this were the ‘other antihypertensive’ group and BBs, which were both associated with increased risk of new-onset treatment for BD. As above, additional analyses indicated that this elevated risk for BBs was restricted to propranolol.

## Discussion

Overall, our findings do not provide support for the repurposing of antihypertensive drugs as treatments for depression. Relative to the comparison group, most classes of antihypertensive were associated with small increased risks of being treated for MDD and a slightly lower risk of being treated for new-onset BD. The main exceptions to this were for patients treated with BBs, which were associated with increased risk of subsequent treatment for both MDD and BD (this is perhaps unsurprising given the widespread use of BBs as anxiolytics).

Our results are consistent with studies suggesting that propranolol may be associated with increased long-term risk of mood disorders (Luijendijk & Koolman, 2012; Verbeek et al.) Additionally, given our study design, our findings are consistent with of the work of Kessing and colleagues (Kessing et al., 2019a, 2019b). Relative to untreated comparison group’s, both Kessing et al and our study had similar results in that prescriptions for AAs were associated with a small increased risk of depression (Kessing et al., 2019b) and a reduced risk of bipolar disorder (Kessing et al., 2019a).

Our design is an advance on previous work (Boal et al., 2016) which did not include a control group. Kessing and colleagues correctly highlight that people with hypertension have a small increased risk of future depression. Their solution was to compare periods where patients had received a cumulative 3 or more prescriptions of AAs relative to a reference period which included people who had only 1 or 2 prescriptions for AAs. While this approach might to some extent address the confounding effects of hypertension, it does also have a potential to add other biases. The comparison group used by Kessing and colleagues included people who only ever received one or two prescriptions of AAs. In our study, those who received an initial treatment with AAs but then swapped to other antihypertensives had greatly elevated risk of depression, suggesting that the control group in the Kessing and colleagues’ study may be confounded by other aspects relating to adherence to medication (Kessing et al., 2019a., 2019b)

Using observational data it is extremely unlikely that any one study design will address all potential biases, and it is necessary to compare across studies and designs, a process termed triangulation (Lawlor, Tilling, & Davey Smith, 2016). On balance, it would appear from the existing literature that AAs are a good candidate for repurposing to treat bipolar disorder. The evidence for repurposing antihypertensive drugs to treat major depressive disorder is weaker. Previously reported protective associations (L. V. Kessing et al., 2019) and the negative associations in our study are weak and occur across different classes of medication. The similarity of associations across multiple classes of medication might reflect non-biological mechanisms and it is well established that patients who are in regular contact with their General Practitioners (for example, for medication reviews or monitoring of blood pressure) are more likely to have psychological problems identified than patients with fewer regular consultations (Bushnell, 2004). Ultimately only a well-designed RCT is likely to resolve these issues but it is unclear at this stage whether the opportunity costs in carrying out such a study are justified.

### Limitations

In 2011, which is within our study period, guidelines on the treatment of hypertension changed and BBs were no longer a preferred initial therapy for hypertension (National Institute for Health and Care Excellence (NICE), 2011). As noted above, BBs such as propranolol are commonly used in primary care settings to treat anxiety (P. E. Hayes & Schulz, 1987), as well as for thyrotoxicosis, and angina. It was not possible for us to identify from prescription records the exact reasons for treatment choices. The observed increased risk of new-onset MDD in this group might therefore reflect the exacerbation of an already-established affective disorder.

The comparison groups clearly differ with respect to cardiovascular history and, despite the cohorts and comparison groups being selected on the basis of their mood disorder histories, they may also differ with respect to subclinical symptoms of anxiety and depression. However, for cohort 1 these differences were likely to be small for all antihypertensive classes, except BBs. In contrast, cohort 2’s risk of mood disorders (relative to their comparison group) continued to increase over time. This could reflect poorer mental health among cohort 2, undiagnosed onset of new mental illnesses among the comparison group, or residual confounding factors. Potentially eligible participants for whom we could not find matches differed from those analysed in that they were older and tended to have poorer health.

New-onset bipolar disorder was primarily identified using PIS records. As such, it was not possible to distinguish between bipolar depressive states and hypomanic or manic states based on the data that we had access to. In addition, given that most cases of bipolar disorder tend to have an onset in early adulthood (many years before treatment for hypertension usually starts), it is not clear that we have accurately captured new-onset bipolar disorder. It is also possible that some of these patients may have received treatment for bipolar disorder earlier in life, during periods (before 1981 for SMR04 and before 2009 for PIS) for which data was not available.

Our study design identified people who *consistently* received antihypertensive monotherapy in order to investigate possible protective effects of antihypertensive treatments in the medium-to long-term. As such, this study was not designed to investigate immediate side effects of antihypertensive treatments. A patient who developed depressive symptoms and received treatment for MDD quickly after their first antihypertensive medication prescription would, by design, be included within cohort 2. However, it is difficult to distinguish such people from those who were selected into cohort 2 due to having prior mental illness. Consequently, for cohort 2, which we considered our secondary analyses, the more interesting aspects of the results are for periods occurring at least 6 months after the initial period of eligibility. A patient who changed their antihypertensive treatments in the first 12 months of treatment, perhaps due to side-effects, would have been included in the ‘other antihypertensive’ group, such that membership of this group may indicate additional health problems beyond hypertension. We also did not have data on whether or not hypertension was being adequately controlled by medication. This could potentially confound the results. Another challenge was the classification of patients treated with multiple antihypertensive treatments. We used a pragmatic measure of whether people had received treatment from at least two antihypertensive classes within the final 3 months of the window used to define eligibility. In theory, this might include people on changing prescription regimes. However, the risks of MDD and BD for people in the polytherapy group were within the range of most monotherapies, suggesting that this was a reasonably robust approach.

Using a comprehensive national-level routine healthcare data linkage approach, we found little evidence to support the idea that antihypertensive medications might be usefully repurposed as treatments for MDD. Tentatively, we conclude that some classes of antihypertensive - and angiotensin antagonists in particular - may offer some protection against BD, but this could be due to selection biases, and will require other study designs to resolve.

## Data Availability

Data cannot be shared publicly because it includes confidential medical records. Researchers can apply for access by submitting an application to the Public Benefit and Privacy Panel (PBPP) at NHS Scotland ISD.

## Acknowledgements

The authors would like to acknowledge the support of the eDRIS Team (National Services Scotland) for their involvement in obtaining approvals, provisioning and linking data and the use of the secure analytical platform within the National Safe Haven.

